# Large Language Models in Cardiology: A Systematic Review

**DOI:** 10.1101/2024.09.01.24312887

**Authors:** Moran Gendler, Girish N Nadkarni, Karin Sudri, Michal Cohen-Shelly, Benjamin S Glicksberg, Orly Efros, Shelly Soffer, Eyal Klang

## Abstract

**Purpose:** This review analyzes the application of large language models (LLMs), in the field of cardiology, with a focus on evaluating their performances across various clinical tasks.

**Methods:** We conducted a systematic literature search on PubMed for studies published up to April 14, 2024. Our search used a wide range of keywords related to LLMs and cardiology to capture various relevant terms. The risk of bias was evaluated using the QUADAS-2 tool.

**Results:** Fifteen studies met the inclusion criteria, categorized into four domains: chronic and progressive cardiac conditions, acute cardiac events, cardiology education, and cardiac monitoring. Six studies addressing chronic conditions demonstrated variability in the accuracy and depth of LLM-generated responses. In acute cardiac scenarios, three articles showed that LLMs provided medical advice with mixed effectiveness, particularly in delivering CPR instructions. Two studies in educational cardiology revealed high accuracy in answering assessment questions and interpreting clinical cases. Finally, four articles on cardiac diagnostics showed that multimodal LLMs displayed a range of capabilities in ECGs interpretation, with some models performing at or exceeding the level of human specialists.

**Conclusion:** LLMs demonstrate considerable potential in the field of cardiology, particularly in educational applications and routine diagnostics. However, their performance remains inconsistent across various clinical scenarios, particularly in acute care settings where precision is critical. Enhancing their accuracy in interpreting real-world complex medical data and emergency response guidance is imperative before integration into clinical practice.

## Introduction

Large language models (LLMs) like OpenAI’s ChatGPT, Google’s Gemini, and Meta’s Llama, are advancing natural language processing (NLP) by generating, understanding, and interpreting text. These models process text to produce coherent responses, understand context, summarize information, and engage in conversations ^1^. Their application in healthcare, particularly in cardiology, offers significant benefits due to their ability to analyze diverse and complex data—from patient records to imaging studies^2–4^.

In cardiology, LLMs are increasingly being used to assist in the management of cardiovascular diseases by organizing and making clinical data more accessible. ^5^. These models can enhance diagnostic accuracy, personalize treatment plans, and identify patterns in large datasets that traditional methods might overlook^6,7^. Additionally, LLMs offer the potential to automate routine documentation, thereby reducing the administrative burden on healthcare providers^8–10^.

However, the integration of LLMs into clinical workflows poses challenges. Effective implementation is crucial to leverage their full potential in improving patient care in cardiology^9,11,12^. This review focuses on the current use of LLMs in cardiology, detailing their potential impact on care and patient outcomes and discussing the barriers to their practical application.

### Overview of Large Language Models in Medical Practice

**Artificial Intelligence (AI)** is a technology designed to perform tasks that typically require human cognition. These include tasks such as decision-making, language comprehension and generation, pattern recognition, and learning from past experiences.

**Deep Learning**, a subset of AI, is modeled after the biological neurons, in processing information through artificial neural networks (ANN) ^13^. These networks consist of “neuron” layers. Each artificial neuron is similar to a logistic regression unit, with inputs coming from the previous layer, and output sent to the next layer.

When physicians analyze patient’s record, they process the information to identify patterns, such as symptoms pointing to a specific diagnosis. Similarly, in deep learning, the input layer may receive patient records or medical images. As the data flows, each layer extracts, and transforms features, allowing the model to recognize complex patterns that might indicate clinical insights. The model’s final output layer then provides the prediction or classification, such as identifying a medical condition. During the training phase, the network adjusts its internal parameters to minimize errors, improving the accuracy of its predictions.

**Natural Language Processing (NLP)**, is a computer field which enables machines to interpret and manage human language. It allows computers to perform tasks such as translating medical texts, detecting the sentiment in clinical notes, or summarizing large documents. This capability enhances the efficiency of information management in clinical practice.

**Attention Mechanisms** represent a major advancement in deep learning models, allowing them to focus on the most relevant parts of input data, akin to how a doctor prioritizes the most critical symptoms during diagnosis. This focus enhances the model’s ability to generate coherent and contextually appropriate responses.

**Transformers**, a powerful class of deep learning models, have revolutionized NLP by processing sequences of text in parallel and using self-attention mechanisms. These mechanisms enable the model to understand the relationships between different parts of the text, significantly improving the speed and accuracy of tasks like automated clinical documentation.

**Large Language Models (LLMs)** are sophisticated AI models designed to process and generate text that closely mimics human writing^14^. Trained on large datasets, LLMs grasp the subtleties of language and context, aiding in tasks such as drafting clinical notes, generating educational materials and supporting medical research. The development of LLMs begins with **pre-training**, where the model learns general language patterns, grammar, and knowledge from a broad corpus of text data. It is followed by fine-tuning, where the pre-trained model is adapted to specific medical domains using specialized datasets, which enhances the model’s relevance in clinical settings ^15^.

The initial input given to an LLM, known as a **prompt**, directs its text generation and sets the direction and tone of the response. Effective prompts are needed for ensuring relevant and accurate outputs.

## Methods

This review was conducted according to the Preferred Reporting Items for Systematic Reviews and Meta-Analyses (PRISMA) guidelines^16^.

### Search Strategy

A literature search was conducted to identify studies on the application of LLMs in cardiology. The search was performed on April 14, 2024, using the PubMed database with a wide range of keywords related to “LLMs” and “Cardiology” to capture various relevant terms. Complete search strategies are detailed in Supplementary Material 1. The study is registered with PROSPERO (CRD42024556397)^17^.

### Study Selection

We included studies that (1) evaluated an application of LLMs in a specific field within cardiology, (2) were published in English, (3) and were peer-reviewed original publications. We excluded non-LLM articles, non-cardiology articles, and non-original articles. Abstracts were also excluded. The search was supplemented by manually reviewing the references of included studies.

Two reviewers (MG, SS) independently screened the titles and abstracts to determine if the studies met the inclusion criteria. Full-text articles were reviewed when the titles met the inclusion criteria or when there was any uncertainty. Disagreements were resolved by a third reviewer (EK).

### Data Extraction

Two independent reviewers (MG, SS) extracted data from the included studies using a standardized data extraction form. Discrepancies were resolved through discussion or consultation with a third reviewer (EK). Extracted information included: study design, sample size, LLM application details (e.g., LLM features examined, assessment method, validation metrics, and reference guidelines used for accuracy comparison), main findings, and limitations.

### Quality Assessment

The quality of the included studies was assessed using an adapted version of the Quality Assessment of Diagnostic Accuracy Studies (QUADAS-2) criteria^18^.

### Data Synthesis

A narrative synthesis of the findings from the included studies was conducted. Due to anticipated heterogeneity in study designs and outcomes, a meta-analysis was not planned. Instead, the focus was on summarizing the applications, benefits, and limitations of LLMs in cardiology as reported in the studies and identifying areas for future research.

## Results

A total of sixteen ^6,11,19–32^ original articles were identified for inclusion. One article was excluded because it was a critical letter pertaining to one of the included studies. Consequently, fifteen articles were selected for review (**Figure 1**).

**Figure 1.**
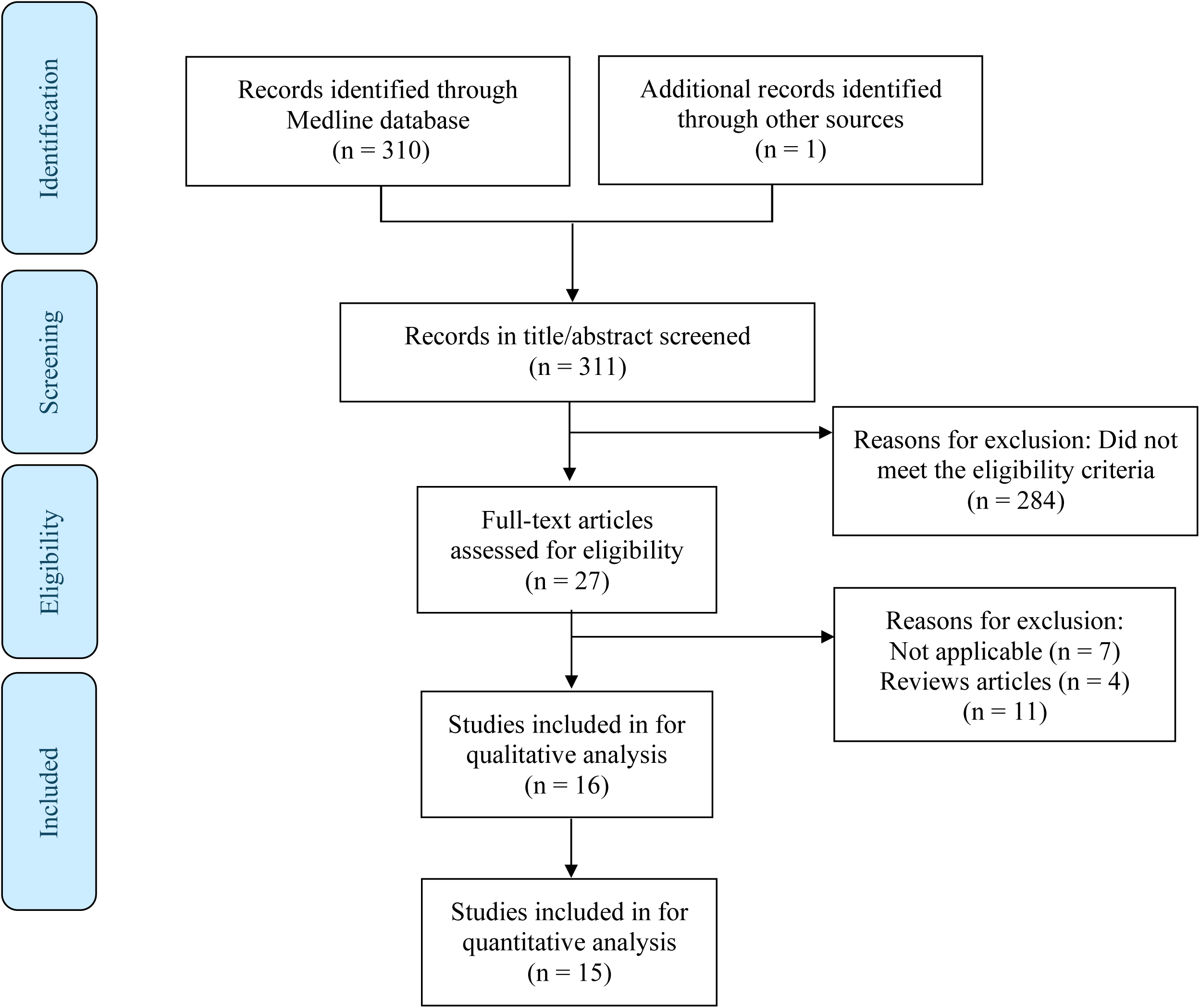
Flow diagram of the search and inclusion process.

These articles were varied in their tasks, and they were organized into four primary categories (Figure 2): 1) Chronic and Progressive Cardiac Conditions; 2) Acute Cardiac Events; 3) Educational Cardiology; 4) Cardiac Diagnostics test.

**Figure 2.**
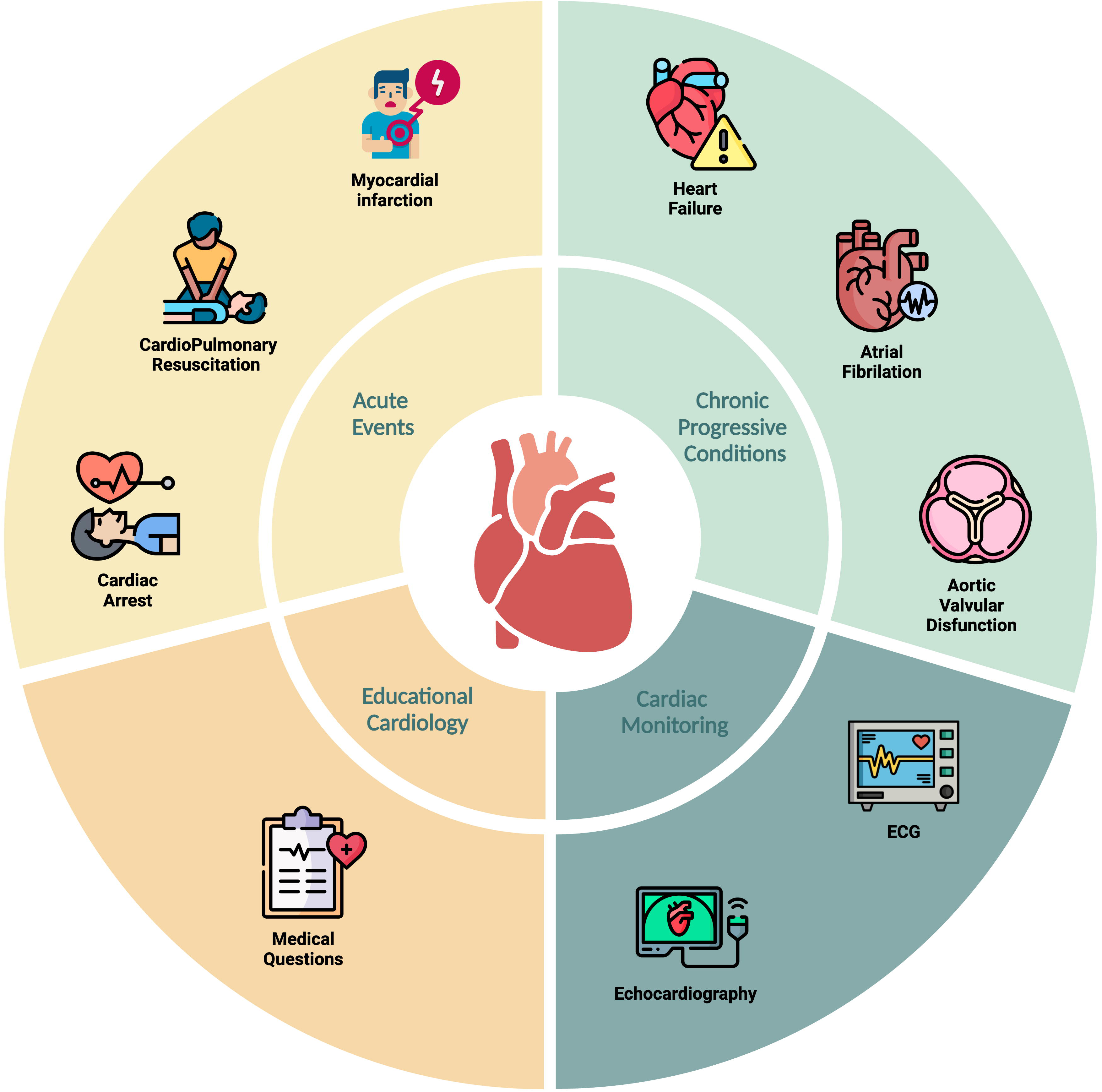
Categorization of Articles into Core Groups with Corresponding Cardiology Subfields.

General details about the included articles, descriptions of their characteristics, main outcomes, and their advantages and limitations are summarized in **Tables 1-4**, respectively. Supplementary Table 1 provides a detailed evaluation of each article using the QUADAS-2 tool.

**Table 1:**
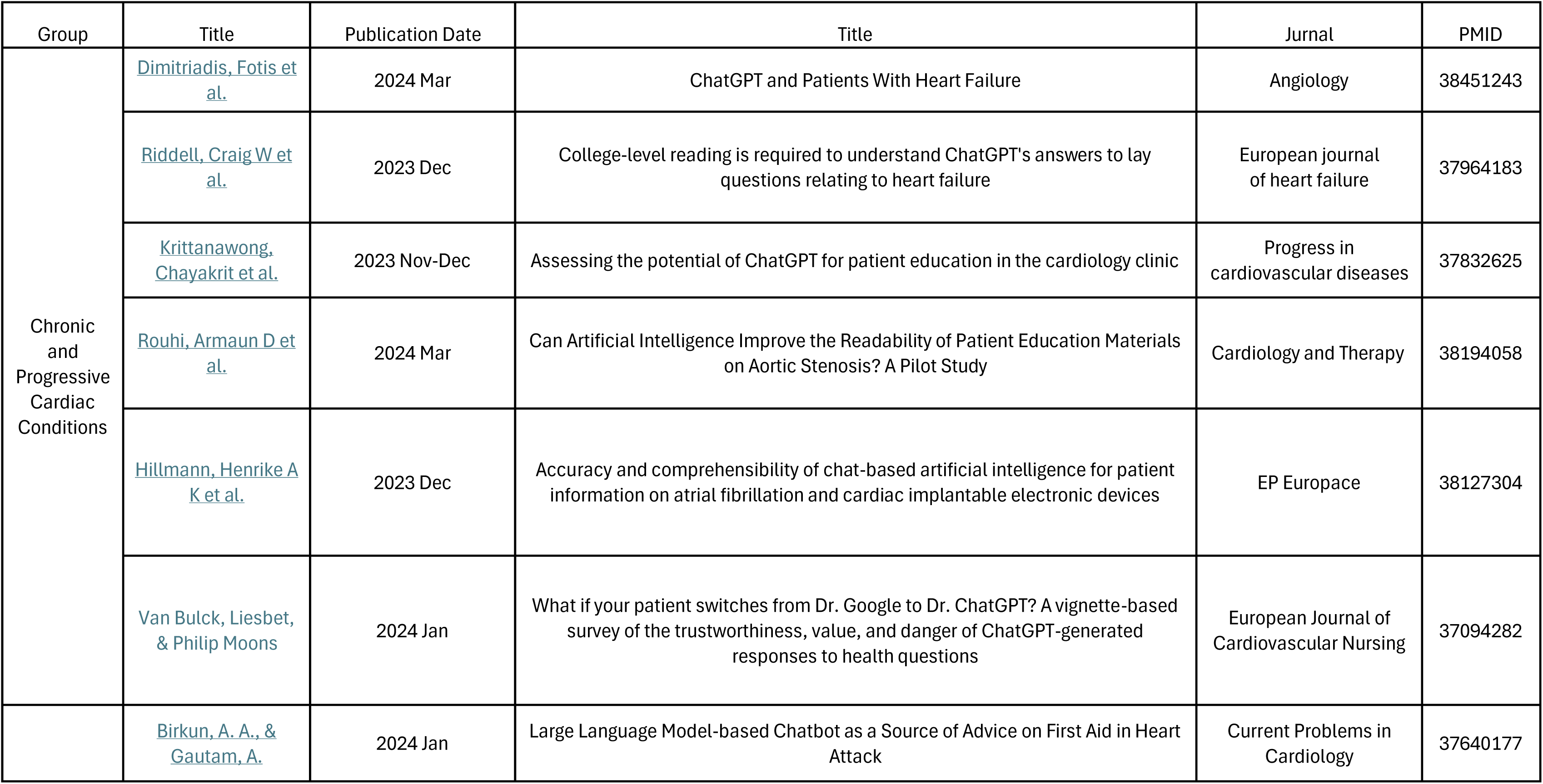
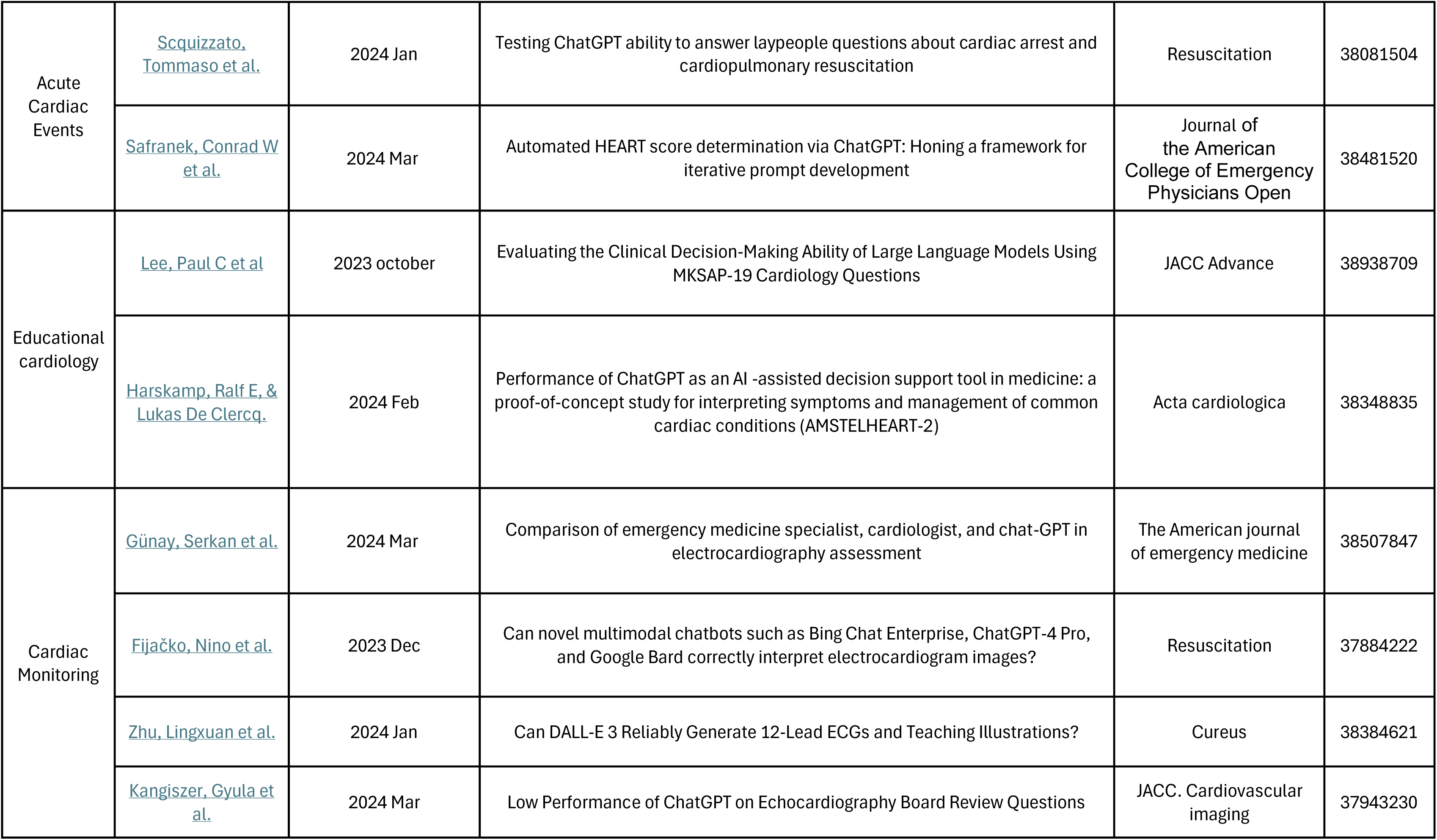
General Details of Studies. The table provides an overview of the studies that have explored. It includes key information such as study titles, authors, publication years, and journal details.

**Table 2:** Study Characteristics. The table summarizes the study characteristics of the studies included in the review. It outlines the basic study parameters such as study design, sample size, population characteristics, and key interventions or technologies evaluated. The table serves as a quick reference to understand the scope and context of each study.

| Group | Title | Tasks | LLM features<br>exmined | Field in<br>Cardiology | LLM type | Dateset | Dateset<br>size (n) | Validation | Validation<br>metrics | Guidelines |
| --- | --- | --- | --- | --- | --- | --- | --- | --- | --- | --- |
| Chronic and<br>Progressive<br>Cardiac<br>Conditions | Dimitriadis, Fotis et al. | Examination the accuracy and reproducibility of ChatGPT in answering questions about knowledge and management of heart failure. | Accuracy | Heart failure | ChatGPT 3.5 | Queries knowledge and menegment of HF. | 47 questions (8 categories) | 2 cardiologies | A study's primary supervisor evaluated researches assesment. | The latest guidelines for HF |
|  | Riddell, Craig W et al. | Examination the readability and broad quality of ChatGPT responses to a set of hypothetical queries from a patient with HF with reduced ejection fraction (HFrEF). | Readability | Heart failure | ChatGPT 4 | Queries knowledge and menegment of HF. | 16 | FRE, SMOG | scoring readability by FRE and SMOG. | American Heart Association<sup>4</sup>and European Society of Cardiology |
|  | Krittanawong, Chayakrit et al. | Examination the appropriateness of the AI model responses to heart failure (HF) related questions. | Reliability | Heart failure | ChatGPT 3.5 | Typical inquiries from patients regarding heart failure and the cardiology clinic. | 20 | Experienced cardiologists and HF specialists | Expert clinical judgment - “reliable but needs explanation”, “reliable without explanation”, or “unreliable”. |  |
|  | Rouhi, Armaun D et al. | Examination the ability of ChatGPT-3.5 and Bard to rewrite patient education materials (PEMs) to meet recommended reading skill levels for patients with AS. | Readability Simplification | Aortic stenosis | Google Bard ChatGPT 3.5 | Patient education materials on AS should address inquiries in simpler language. | 21 | FRE, FKGL, GFI, SMOGI | FRE score ranking 0-100 GFI, and SMOGI range from 0 to 18, 0 to 20, and 5 to 18, respectively, and each score indicates the number of years of education necessary to understand the assessed reading material. | Professional cardiothoracic surgical society and academic institutions in the USA |
|  | Hillmann, Henrike A K et al. | Examination the different aspects of responses given by different NLPs on questions about atrial fibrillation (AF) and clinical implantable electronic devices (CIED). | Readability Appropriateness Comprehensibility Missing of content Confabulation Clinical decisions | Atrial fibrillation (AF) clinical implantable electronic devices (CIED) | Google Bard Bing Chat ChatGPT 4 | Typical inquiries from patients regarding AF or CIED. | 50 (25 on AF, 25 on CIED) | 3 experienced electrophysiologists | Expert rating |  |
|  | Van Bulck, Liesbet, & | Examination the trustworthiness, value, and danger | Reliability Confabulation | Chronic heart disease (CHD) Atrial | ChatGPT 3 | Typical inquiries from virtual | 4 prompts | 20 (19 nurses, 1 dietician) | Each expert rated the case on a scale |  |
|  | Philip Moons | of information provided by ChatGPT on virtual prompts by patients. |  | fibrillation (AF)<br>Heart failure (HF)<br>Cholesterol (Chol) |  | patients regarding CHD, AF, HF and Chol. |  |  | from 1 to 10. Comparing the valuability to Google searching. |  |
| Acute Cardiac Events | Birkun, A. A., & Gautam, A. | Examination the ability of chatbot's to guide first aid for heart attacks. | Adherence to guidelines | Myocardial infarction (MI) | Bing Chat | Typical inquiries from patients regarding MI menegmen t. | 60 (20 for a group) | Researches | Using a checklist with ratings ( True, Partially True, or Not True) of content satisfaction. | International Federation of Red Cross and Red Crescent Societies' International First Aid, Resuscitation, and Education Guidelines 2020. |
|  | Scquizzato ,Tommaso et al. | Examination the precision of ChatGPT's responses to public inquiries regarding cardiac arrest and CPR | Accuracy Readability | Cardiac arrest CPR | ChatGPT 3.5 | Typical inquiries from patients regarding different aspects of cardiac arrest. | 40 | 2 groups:<br>Group 1 - professional s with expertise in out-of-hospital cardiac arrest (OHCA), including 8 doctors, 5 nurses, and a psychologist.<br>Group 2 consisted of laypeople (survivors, first responders, and family members affiliated with Peer Support Group). | 1 (poor) to 5 (excellent) for each feature exmined. | Sudden Cardiac Arrest UK focusing cardiac arrest and CPR |
|  | Safranek, Conrad W et al. | Examination the ability of ChatGPT to analyze and extract medical information from patient notes and determine the HEART scores in chest pain evaluation. | Extraction information Adherence to guidelines | Educational cardiology | ChatGTP 3.5<br>ChatGPT 4 | Medical notes of patients | 24 | Researchers | Comparison of researchers calculation | HEART score based on History, ECG, Age, Risk factors, and the Troponin risk algorithm |
| Educational cardiology | Harskamp, Ralf E, & Lukas De Clercq. | Examination the accuracy of ChatGPT's recommendations on medical questions related to common cardiac symptoms or conditions. | Accuracy | Educational cardiology | ChatGTP 3.5<br>ChatGPT 4 | Multiple questions Clinical cases | 50<br>20 | Medical expert who developed the quiz | Trivia questions - Comparison Clinical cases- expert opinion and clinical course | Trivia questions - based on quizzes for medical professionals. Clinical cases - A random sampling of clinical cases that presented to a community health centre in Amsterdam, The Netherlands. |
|  | Lee, Paul C et al | Assessing the performance of two LLMs and physicians in achieving a passing score of 50% using MKSAP- | Accuracy | Educational cardiology | ChatGPT 3.5<br>ChatGPT 4<br>PubMedGPT | Clinical cases | 120 | physicians | Medical Knowledge Self-Assessment Program (MKSAP-19) | Medical Knowledge Self-Assessment Program (MKSAP-19) |
|  |  | 19's cardiology questions |  |  |  |  |  |  |  |  |
| Cardiac Monitoring | Günay, Serkan et al. | Examination GPT-4 diagnoses using ECG data and compare it to emergency doctors and cardiologists. | Accuracy | ECG | ChatGPT 4 | Multiple questions with ECG cases | 40 (20 easy and 20 challenging) | 12 emergency medicine specialists and 12 cardiology specialist | Comparison | "150 ECG Cases" book |
|  | Fijačko, Nino et al. | Examination the accuracy of three large language models toninterpret ECG images. | Accuracy | ECG | Google Bard<br>Bing Chat<br>ChatGPT 4 | Multiple questions | 81 (27 for each Chatbot) | AHA ACLS exam answers | Comparison | American Heart Association (AHA) Advanced Cardiovascular Life Support (ACLS) multi-choice question exams |
|  | Zhu, Lingxuan et al. | Examination whether DALL-E 3 could extend the material for clinical research and medical education. | Creating medical education tools | ECG CPR | ChatGPT 4<br>DALL-E 3 | Descriptions of 12 leads ECG and CPR | Not relevant | Researchers | Comparison |  |
|  | Kangiszer, Gyula et al. | Examination the ability of ChatGPT Plus to answer correctly echocardiography board review questions and also to provide explanations that reflect standards of practice. | Accuracy | Echocardiography | ChatGPT 4 | Multiple questions (with and without forced justification)<br>Open ended questions | 150 | Answers of Klein's book | Comparison (binarized 1 for correct, 2 for incorrect) | Klein's Clinical Echocardiography Review: A Self-Assessment Tool textbook |

**Table 3:** Outcomes of LLM Applications. The table presents the outcomes and findings of the studies of LLMs in cardiology. It covers various aspects such as accuracy, reliability, clinical relevance, and the potential impact on patient care.

| Group | Title | LLM features examined | Qualitative | Quantitative | Regarding to the features |
| --- | --- | --- | --- | --- | --- |
| Chronic and Progressive Cardiac Conditions | Dimitriadis, Fotis et al. | Accuracy Quality | ChatGPT was able to provide correct answers to most questions in a simple and explanatory way.<br>SGLT2s were not listed as a treatment option. | 43 (differently in 4 of 47 questions). | **Accuracy-**<br>signs and symptoms, 7/7 right answers<br>Lifestyle, 9/11 right answers<br>Treatment, 6/9 right answers<br>Living with HF 8/9 right answers |
|  | Riddell, Craig W et al. | Readability Quality | **Quality -**<br>SGLT2s were not listed as a treatment option.<br>Long detailed answers with lack of emphasis on the key learning points.<br>Frequent usage of jargon, sometimes without explanation.<br>Overall, answers to the same questions were judged as having high consistency, with only minor variations between separate chatbot sessions. | **Correct responses-**<br>6% of answers - college graduate reading level<br>71% of answers - college reading level<br>23% of answers - lower than college reading level<br>4% of answers - grade 8-9 reading level | **Readability-**<br>FRE (31.5,30.1,32.9) collage reading<br>SMOG (16,16.3,15.6) undergraduate reading |
|  | Krittanawong, Chayakrit et al. | Reliability |  |  | **Reliability**<br>8 of 20 questions (40%) were graded as reliable without explanation<br>8 of 20 questions (40%) were graded as reliable with explanation. |
|  |  |  |  |  | **Appropriateness-**<br>4 responses (20%) were graded as inappropriate in both contexts. |
| | <u>Rouhi, Armaun D et al.</u> | Readability<br>Accuracy<br>Rewriting | Neither Platform:<br>Generated PEMs above the recommended 6th-grade reading level.<br>ChatGPT-3.5 demonstrated better post-conversion readability scores, percentage change, and conversion time (all $p < 0.001$). | | **Readability-**<br>original materials - 10th-12th grade reading level.<br>ChatGPT-3.5: 6th-7th grade level grade reading level ($p < 0.001$)<br>Bard: 8th-9th grade reading level ($p < 0.001$) (except for SMOGI $P = 0.729$) |
|  | <u>Hillmann, Henrike A K et al.</u> | Readability<br>Appropriateness<br>Comprehensibility<br>Missing of content<br>Confabulation<br>Clinical decisions |  |  | **Readability:**<br>Varied across different NLPCs<br>**Appropriateness:**<br>AF Responses: GB (52%), BC (60%), CGP (84%)<br>CIED Responses: GB (16%), BC (72%), CGP (88%)<br>**Missing Aspects:**<br>AF: GB (52%), BC (60%), CGP (24%)<br>CIEDs: GB (92%), BC (88%), CGP (52%)<br>**Comprehensibility:**<br>AF Responses: GB (96%), BC (88%), CGP (92%)<br>CIED Responses: GB (92%), BC (88%), CGP (100%):<br>**Missing relevant information:**<br>AF Responses: GB (52%), BC (60%), CGP (24%) .<br>CIED Responses: GB (92%), BC (88%), 52 (100%) |
|  | <u>Van Bulck, Liesbet, & Philip Moons</u> | Trustworthiness<br>Value<br>Danger of information | Against Google's information, 40% of experts found ChatGPT more valuable, 45% equally valuable, and 15% less valuable. |  | Trustworthiness - 7<br>valuable - 7.5<br>Danger of information - 3 |
| Acute Cardiac Events | <u>Birkun, A. A., & Gautam, A.</u> | Accuracy<br>Readability<br>Adherence to guidelines | Chatbot responses had similar length for the 3 studied countries.<br>whereas the responses always suggested to prompt the person to chew a dose of acetylsalicylic acid, they never mentioned the recommended dose of the drug (150-300 mg). | **guidelines inconsistent responses -**<br>Gambia - 25.0% (n = 5)<br>India - 45.0% (n = 9)<br>USA- 25.0% (n = 5) | **Readability-**<br>Gambia and USA - 12 education grade<br>India - 10th grade<br>p<=0.008<br>**Adherence to guidelines**<br>Measured by the number of source web articles cited in a single chatbot<br>**Adherence to guidelines**<br>Gambia 2.0 [1.0-2.0]<br>India 1.0 [1.0-1.0]<br>USA 2.0 [1.0-2.0]<br>Gambia 1-3<br>India 1-5<br>USA 1-3 |
|  | <u>Scquizzato, Tommaso et al.</u> | Accuracy<br>Readability | Professionals rated it lower at 4.0 (±0.5) compared to laypeople's 4.6 (±0.7); with a significance level of p = 0.02.<br>**CPR-Related Answers:**<br>Scored consistently lower across all parameters by both professionals and laypeople. | ChatGPT's responses were given a positive average score of 4.3 (±0.7) by 14 professionals and 16 laypeople. | **Accuracy** - 4.0 (±0.6).<br>**Clarity** - 4.4 (±0.6) P=0.19<br>**Relevance** - 4.3 (±0.6) P=0.09<br>**Comprehensiveness** - 4.2 (±0.7) P=0.02<br>**CPR-**<br>Median Flesch reading ease score of 34 [Interquartile Range (IQR) 26–42]. |
|  | Safraneck, Conrad W et al. | Extraction information<br>Adherence to guidelines | In both models there was a decrease in the rate of responses with erroneous, non-numerical subscore answers. | Corect prediction regarding to HEART score<br>GPT-3.5 - 81.5% (71.7%–88.4%)<br>GPT-4 - 100% (96.3%–100%) | **Extraction information**<br>Non numerical subscore for synthetic patient notes- Chat 3.5 5.7% (3.6%–8.9%)<br>GPT-4 0.3% (0.1%–1.9%)<br>Numerical subscore for synthetic patient notes- a mean error of:<br>GPT-3.5 0.42 (0.33–0.50)<br>GPT-4 0.42 (0.36–0.48) |
| Educational cardiology | <u>Lee, Paul C et al</u> | Accuracy | | ChatGPT 3.5 met the passing criteria (50%), answering 66 questions correctly (55%), numerically but not statistically lower than physicians (60%, $P = 0.43$). PubMedGPT scoring 27%, significantly lower than physicians ($P < 0.001$) and ChatGPT-3.5 ($P < 0.001$). ChatGPT-4 outperformed average MKSAP-19 users (80% vs 60%, $P = 0.0004$), matching the first author, a seasoned cardiologist (80%). | |
| | <u>Harskamp, Ralf E, & Lukas De Clercq.</u> | Accuracy | In 20 case vignettes, ChatGPT's response matched the actual advice given in 85% of cases.<br>All straightforward patient-to-physician questions were answered correctly (10/10).<br>In complex cases where general practitioners asked cardiologists for assistance, ChatGPT was correct in 70% of cases. | ChatGPT's latest version correctly answered 46/50 92% of trivia questions. | **Cardiovascular tricia questions**<br>**Accuracy-**<br>ChatGPT's latest version improves the accuracy compared with an earlier version (92% vs 74%, $p=0.031$)<br>Variation in accuracy-Coronary artery disease (10/10)<br>Pulmonary and venous thrombotic embolism (10/10)<br>Atrial fibrillation (9/10)<br>Heart failure (9/10)<br>Cardiovascular risk management (8/10). |
| Cardiac Monitoring | <u>Günay, Serkan et al.</u> | Accuracy | | GPT-4 -<br>an average of 19.50 ($\pm 0.52$) correct answers in everyday ECG questions<br>an average of 16.83 ($\pm 0.94$) correct answers in more challenging ECG questions<br>an average of 36.33 ($\pm 0.98$) | **Accuracy-**<br>Chat-GPT was found to be more accurate and have superior performance compared to the other groups ($p < 0.001$, $p = 0.001$).<br>In the more challenging ECG questions, Chat-GPT outperformed the emergency medicine specialists ($p < 0.001$), but no significant statistical difference was |
| | | | | correct answers in total ECG questions. | found between it and the cardiology specialists ($p = 0.190$). |
|  | Fijačko, Nino et al. | Accuracy | ChatGPT-4 Plus was the most successful chatbot<br>In interpretation of the ECG images, all the chatbots were inconsistent in answering when questions were repeated. | The overall level of correctness for interpretation of the ECG images was-Bing Chat Enterprise- 67% (95% CI: 58.2–77.7%)<br>ChatGPT-4 Plus -78.9% (95% CI: 74.5– 83.3%)<br>Google Bard - 80.9% (95% CI: 73.4–88.3%) | **Accuracy-**<br>ChatGPT-4 Plus, correctly interpreting almost two-thirds of ECG images (17/27; 63.0%)<br>Google Bard, correctly interpreting ECG images almost half of the time (13/27; 48.2%)<br>Bing Chat Enterprise, correctly interpreting ECG images less than a quarter of the time (6/27; 22.2%). |
|  | Zhu, Lingxuan et al. | Creating medical education tools | **ECG-**<br>DALL-E 3 was able to produce an image containing some key elements of an ECG tracing, the waveform itself did not reflect a physiologically accurate or interpretable ECG. The heart rate was inaccurate, and important features like T waves were missing.<br>**CPR-** illustrations were clear and depicted medical procedures, however they contain some spelling errors. |  |  |
|  | <u>Kangiszer, Gyula et al.</u> | Accuracy<br>Providing explanation | ChatGPT 4 provided a single option as an answer to 141 questions across the 3 question formats.<br>ChatGPT demonstrated the most accuracy in answering questions derived in MC-J format. | The mean cosine similarity for explanations of the correct answers asked in MC-NJ format compared with Klein was 0.93 preprocessing and 0.94 postprocessing. | **Accuracy-**<br>For the OE, MC-NJ, and MC-J formats, 47.3%, 53.3%, and 55.3% questions were correctly answered.<br>**Providing explanation-**<br>Fact-based from correct answers-OE -81.6% (fact based), 12.6% (clinical vignette), and 5.6% (formula based).<br>MC-NJ 81.2%(fact based), 12.5%(clinical vignette), and 6.2% (formula based).<br>MC-J 81.9%(fact based), 12.0%(clinical vignette), and 6.0% (formula based). |

**Table 4:** Strengths and Limitations. The table outlines the strengths and limitations of each study included in the review. It captures the critical evaluation of each study’s methodology, results, and applicability to real-world clinical settings. The table helps identify the robustness of the studies and areas where further research is needed.

| Group | Title | LLM Advantages | LLM Disadvantages | Limitations | Conclusion |
| --- | --- | --- | --- | --- | --- |
| Chronic and Progressive Cardiac Conditions | <u>Dimitriadis, Fotis et al.</u> | <u>Good Performance:</u> The cardiologist deemed ChatGPT's performance as very good.<br><u>Free Access:</u> The public can use ChatGPT at no cost, benefiting individuals with limited financial resources.<br><u>Efficient Data Analysis:</u> ChatGPT can analyze extensive data and provide comprehensible replies.<br><u>Patient-Friendly Language:</u> ChatGPT provides answers in a simple, non-medical manner.<br><u>Enhanced Accessibility:</u> Real-time information availability can alleviate anxiety for patients. | <u>Ensuring data transparency and accountability:</u> crucial for user security. Users need to fully comprehend how their data is utilized and who can access it.<br><u>Risk for Undiagnosed Users:</u> Individuals without prior diagnosis may be exposed to possible risks by relying solely on ChatGPT for web-based diagnostic purposes.<br><u>Content Moderation Effort:</u> Significant effort to prevent offensive content, but effectiveness remains uncertain.<br><u>Long Responses:</u> Tends to generate lengthy replies without references.<br><u>Lack of Real Patients:</u> The study lacks actual patient involvement, affecting patient understanding. | <u>Trustworthiness Assessment:</u> ChatGPT lacks the ability to discern trustworthy information from falsehoods due to reliance on web-based content.<br><u>Lack of Real-Time Information:</u> ChatGPT's training data is up to September 2021, limiting access to real-time events. | ChatGPT's accuracy in responding to heart failure management questions is promising, but further investigation is needed to assess its reliability. |
|  | <u>Riddell, Craig W et al.</u> | High level of confidence in the validity -by separate measures of readability across three separate sets of responses to the questions. <u>Hallucination</u> were not detected<br><u>Minimal key detail omissions</u> were revealed, predominantly related to recently approved HF medications | No assessment to other sources of reading complexity such as use of jargon. |  | ChatGPT has potential as a patient medical support, but further research is needed to fully evaluate it. |
|  | <u>Krittanawong, Chayakrit et al.</u> | ChatGPT model provided primarily reliable answers to several commonly asked questions related to heart failure |  | <u>Trustworthiness Assessment:</u> ChatGPT lacks the ability to discern trustworthy information from falsehoods due to reliance on web-based content. | ChatGPT may be useful to expedite workflows in HF and cardiology clinics, but AI alone is not sufficient to answer all HF questions. . |
|  | <u>Rouhi, Armaun D et al.</u> | The analysis did not identify any factual errors in the AI-generated medical content. | The AI dialogue platforms only received one prompt in this study.<br>Better phrases may exist to query LLMs regarding PEM readability improvement<br>PEMs from countries outside the USA were not analyzed | The study utilized free versions of ChatGPT-3.5 and Bard, whereas paid versions of these same AI platforms may include higher-level capabilities and functioning. | AI dialogue platforms, particularly ChatGPT-3.5, can enhance the readability of Patient Education Materials (PEMs) for patients with Aortic Stenosis (AS), although they may not fully meet recommended reading skill levels. This highlights the potential of these platforms as tools to strengthen cardiac health literacy in the future. |
|  | <u>Hillmann, Henrike A K et al.</u> | Optimized Access: NLPs enhance medical information accessibility.<br>Understandable Advice: Responses are clear across different NLPs.<br>Patient Education: NLPs | Incomplete Advice: Key aspects often missing, especially with Google Bard or BC.<br>Misstatements: Experts noted minor but significant errors in responses. | <u>Subjective Questions:</u> Lack of standardized questionnaire; limited expert evaluation.<br><u>Expert Bias:</u> Results may not reflect patient comprehension.<br><u>Outdated Training:</u> Chatbot | NLPC Responses are mostly easy to understand with varying readability. However, it should be used with caution to gather medical information about AF and CIEDs |
|  |  | may boost learning about personal health. |  | knowledge limited to data up to a certain date. |  |
|  | <u>Van Bulck, Liesbet, & Philip Moons</u> | Value and trustworthiness of responses rated positively. | <u>Common negative feedback:</u> missing information, vagueness, and lack of patient-centered writing. No medical doctors or patient representatives included. | <u>Lack of Real-Time Information:</u> ChatGPT's training data is up to September 2021, limiting access to real-time events. Based on GPT-3, slight wording adjustments yield <u>different answers</u>, potentially misleading patients. <u>Risk of inaccurate content</u> and hallucination. | ChatGPT responses are trustworthy and valuable but must be interpreted cautiously due to limitations. |
| Acute Cardiac Events | <u>Birkun, A. A., & Gautam, A.</u> | Value and trustworthiness of responses rated positively. | <u>Low Overall Quality:</u> Despite some instructions aligning with guidelines, the chatbot's guidance quality is low. <u>Omitted Directions:</u> Important first aid instructions are consistently missed, which could improve outcomes in real myocardial ischemia cases. <u>Inaccurate/Incomplete Instructions:</u> Relevant advice is often wrong or incomplete, such as incorrect EMS numbers or inadequate CPR criteria. <u>Excessive Directives:</u> Responses include unnecessary directives that could distract or harm during emergencies. | <u>Under Development:</u> The new Bing chatbot is available to use but is still being developed. <u>Web Content Influence:</u> Changes in web content could affect the chatbot's responses. <u>Inconsistent Content:</u> The number and accuracy of instructions vary, affecting the reliability of the guidance. <u>Poor Source Quality:</u> The chatbot's advice quality is impacted by the inaccuracies and omissions in the source web articles. | Due to subpar response quality, the chatbot is not advised as a reliable first aid information source. |
|  |  |  | <u>No VPN Use:</u> The study didn't use a VPN, which may have limited the simulation of searches from specific network addresses in the Gambia.<br><u>Geographical Inconsistency:</u> The new Bing's responses were of variable and generally low quality, failing to adapt to the search region. |  |  |
|  | <u>Scquizzato, Tommaso et al.</u> | Value and trustworthiness of responses of ChatGPT rated positively both professionals and laypeople. | SCA UK's members are usually younger than the average population of cardiac arrest survivors, are more frequently fitted with an ICD | <u>Inconsistent Content:</u> The number and accuracy of instructions vary, affecting the reliability of the guidance.<br><u>Lack of Real-Time Information:</u> ChatGPT's training data is up to September 2021, limiting access to real-time events. | ChatGPT provided largely accurate answers, except for CPR-related ones. Robust monitoring measures are needed for healthcare-related content generated by these systems. |
|  | <u>Safranek, Conrad W et al.</u> | The framework prompt improvement for automated HEART score determination across a limited set of synthetic patient notes. | Using synthetic medical notes, due to privacy issues. Creating significantly shorter clinical notes than the real world patient's complied clinical notes. | Inaccurate/Incomplete Instructions: Relevant advice is often wrong or incomplete | The study group managed to develop a framework for iterative prompt design in clinical settings. While the results show promise for LLMs, it's crucial to apply this to large-scale clinical data with proper data privacy measures. |
| Educational cardiology | <u>Lee, Paul C et al</u> | GPT-4's impressive performance on the MKSAP-19 cardiology questions indicates its potential ability to process medical information, understand complex medical contexts, and provide evidence-based recommendations. | "Overconfidence, where the model expresses certainty when uncertainty should be acknowledged<br>Incorrect numerical computations<br>Inability to interpret images<br>Hallucination" | * PubMedGPT performed poorly, potentially due to the difference in neural network parameters (ChatGPT: 175 billion, PubMedGPT: 2.7 billion).<br>* E12The exam questions may not reflect real-life clinical challenges. | Integrating LLM into clinical settings has the potential to enhance efficiency, streamline workflows, and ultimately contribute to improved patient outcomes. However, it is crucial to remain vigilant about their limitations and to continue refining their capabilities to ensure their safe and effective use in medical practice. |
|  | <u>Harskamp, Ralf E, & Lukas De Clercq.</u> | Relaying on multiple approaches with variable complexity.<br>Cases vignettes provides a <u>good simulation</u> of how ChatGPT would preform in real life.<br>ChatGPT <u>answered well</u> to multiple questions and relative stright forward medical questions.<br>ChatGPT provided <u>consist</u> answers. | ChatGPT was not fully adressed with complex questions, that need a second consultation. | <u>No transparency</u>- ChatGPT can't provide <u>references</u> to the provided answers. | ChatGPT has potential as an AI-assisted decision support tool in medicine, particularly for straightforward, low-complex medical questions, but further research is needed to fully evaluate its potential. |
| Cardiac Monitoring | <u>Günay, Serkan et al.</u> | ChatGPT showed better accuracy than the other two groups in everyday ECG questions. | GPT-4 may need further improvement and refinement in its understanding and recognition of wide QRS tachycardias. | Absence of visual capacity - the ECG images were described. The evaluation of the Chat is based on the discription. | GPT-4 outperforms emergency doctors in all ECG tests and does better than cardiologists in common ones. However, its performance aligned closely with that of the cardiologists as the difficulty of the questions increased. |
|  | <u>Fijačko, Nino et al.</u> |  |  | the evaluation of the Chat is based on the discription and not on the image.<br><u>Inconsistent Content</u>: The number and accuracy of instructions vary, affecting the reliability of the guidance. | ECG interpretation might be aided by more adept artificial intelligence tools in the near future. |
|  | <u>Zhu, Lingxuan et al.</u> | Serves as a promising tool for simple medical illustrations. Depicting core elements of medical procedures successfully | The ECG it drew could not be considered a standard 12-lead ECG, as it failed to produce obvious T waves and contained several interfering waves.<br>No quantitative scoring of output quality was given. |  | the AI does not yet have a robust enough understanding of ECG physiology and waveform characteristics to produce valid examples. |
|  | <u>Kangiszer, Gyula et al.</u> | ChatGPT provide trainees with a more insightful understanding of the subject and the ability to adapt diction to best suit learners depending on the context of information. | ChatGPT4 demonstrated low performance on echocardiography board exam questions. | There is no comparison the performance of ChatGPT with human trainees. | ChatGPT has potential augment clinical knowledge and prepare clinicians for cardiovascular board certification exams, but further research is needed to fully evaluate it. |

### Chronic and Progressive Cardiac Conditions

Six studies ^19–24^ assessed the accuracy and reliability of LLMs in managing long-term heart conditions like heart failure and valvular defects.

Dimitriadis et al. found that ChatGPT accurately addressed 7 out of 8 frequently asked patient questions about heart failure ^19^. Although the responses with lower accuracy still provided correct information, it was deemed insufficient. This evaluation, conducted by cardiologists using the latest heart failure guidelines, highlighted ChatGPT’s performance.

Riddell et al.^20^ focused on the readability of ChatGPT’s responses, finding that 71% were at a college reading level. In contrast, Rouhi et al. ^22^ found that ChatGPT 3.5 and Google Bard reduced the readability of educational materials on aortic stenosis to a 6th-7th grade level and an 8th-9th grade level, respectively.

### Acute Cardiac Events

Three^25–27^ studies evaluated the effectiveness of AI chatbots in providing medical advice on acute cardiac event. Scquizzato et al.^26^ assessed ChatGPT’s responses to cardiac arrest and cardiopulmonary resuscitation (CPR) questions from laypeople. They found that the answers were overall well received with an average rating of 4.3 out of 5. Clarity (4.4), relevance (4.3), accuracy (4.0), and comprehensiveness (4.2) were highly rated, but CPR-related answers scored lower, especially among professionals.

Birkun et al.^25^evaluated Bing chatbot’s first aid advice for heart attacks in The Gambia, India, and the USA. While the chatbot provided general guidance, it often missed critical life-saving instructions ^27^, such as taking antianginal medication or initiating CPR for unresponsive individuals. The mean percentage of responses fully aligned with the checklist criteria ranged from 7.3% (India) to 16.8% (USA). Furthermore, a significant portion of the responses included inconsistent directives, with 25% for The Gambia and the USA, and 45% for India.

In a proof-of-concept study by Safranek et al.^27^, ChatGPT was evaluated for its ability to determine the HEART score (History, ECG, Age, Risk factors, Troponin risk algorithm) in chest pain evaluation. The accuracy of numerical responses for HEART subscores improved significantly for ChatGPT-4, with the mean error decreasing from 0.16 to 0.10 points. The final prompt versions reduced the rate of non-numerical responses for ChatGPT-3.5 from 18.7% to 6.7% and for ChatGPT-4 from 5.7% to 0.3%, demonstrating the potential for iterative prompt design to enhance clinical decision support tools.

### Educational Cardiology

Two studies^6,28^ assessed the accuracy of ChatGPT’s responses to clinical cardiac questions. Harskamp and De Clercq^28^ tested ChatGPT’s cardiovascular knowledge and clinical case vignettes. ChatGPT-4 correctly answered 92% of assessment questions and matched 85% of the actual medical advice in 20 clinical cases. Straightforward questions were answered correctly 100% of the time, while complex cases requiring specialist input had a 70% accuracy rate. The latest model version showed significant improvement over an older version, with accuracy increasing from 74% to 92% for assessment questions and from 50% to 70% for complex cases.

### Cardiac Diagnostics test

Four studies^11,29–31^ reviewed the performance of LLMs in interpreting cardiac diagnostics test. Fijačko et al.^30^ assessed three multimodal chatbots—Bing Chat Enterprise, ChatGPT-4 Plus, and Google Bard—for interpreting ECG images. ChatGPT-4 Plus was the most accurate, interpreting 63.0% correctly, followed by Google Bard at 48.2% and Bing Chat Enterprise at 22.2%. ChatGPT-4 Plus always provided an interpretation, whereas Bing Chat Enterprise and Google Bard failed in 11.1% and 3.7% of cases, respectively. Google Bard had the highest overall Level of Correctness (LOC) at 80.9%, followed by ChatGPT-4 Plus at 78.9%, and Bing Chat Enterprise at 67%.

Günay et al.^29^ compared GPT-4’s diagnostic accuracy in ECG evaluations with that of emergency medicine specialists and cardiologists. ChatGPT-4 outperformed both groups, with an average of 19.5 correct answers out of 20, compared to 16.75 and 17.08 for emergency medicine specialists and cardiologists, respectively. ChatGPT-4 was particularly successful in challenging questions compared to emergency medicine specialists but showed no significant difference from cardiologists.

Zhu et al.^31^ explored DALL-E 3, integrated with ChatGPT, for generating 12-lead ECGs and medical illustrations. DALL-E 3 produced basic ECGs but lacked accuracy in depicting T waves and heart rate. However, it excelled in creating clear CPR instructional illustrations.

Kangiszer et al.^11^ evaluated ChatGPT-4’s performance on echocardiography with 150 questions in various formats. ChatGPT-4 correctly answered 47.3% of open-ended questions, 53.3% of multiple-choice without forced justification, and 55.3% of multiple-choice with forced justification. Fact-based questions had the highest accuracy, with a cosine similarity of 0.94 compared to standard textbook answers.

Figure 3 outlines the three primary aspects explored in the articles, including the reliability of LLMs, user interaction, and their specific applications in cardiology.

**Figure 3.**
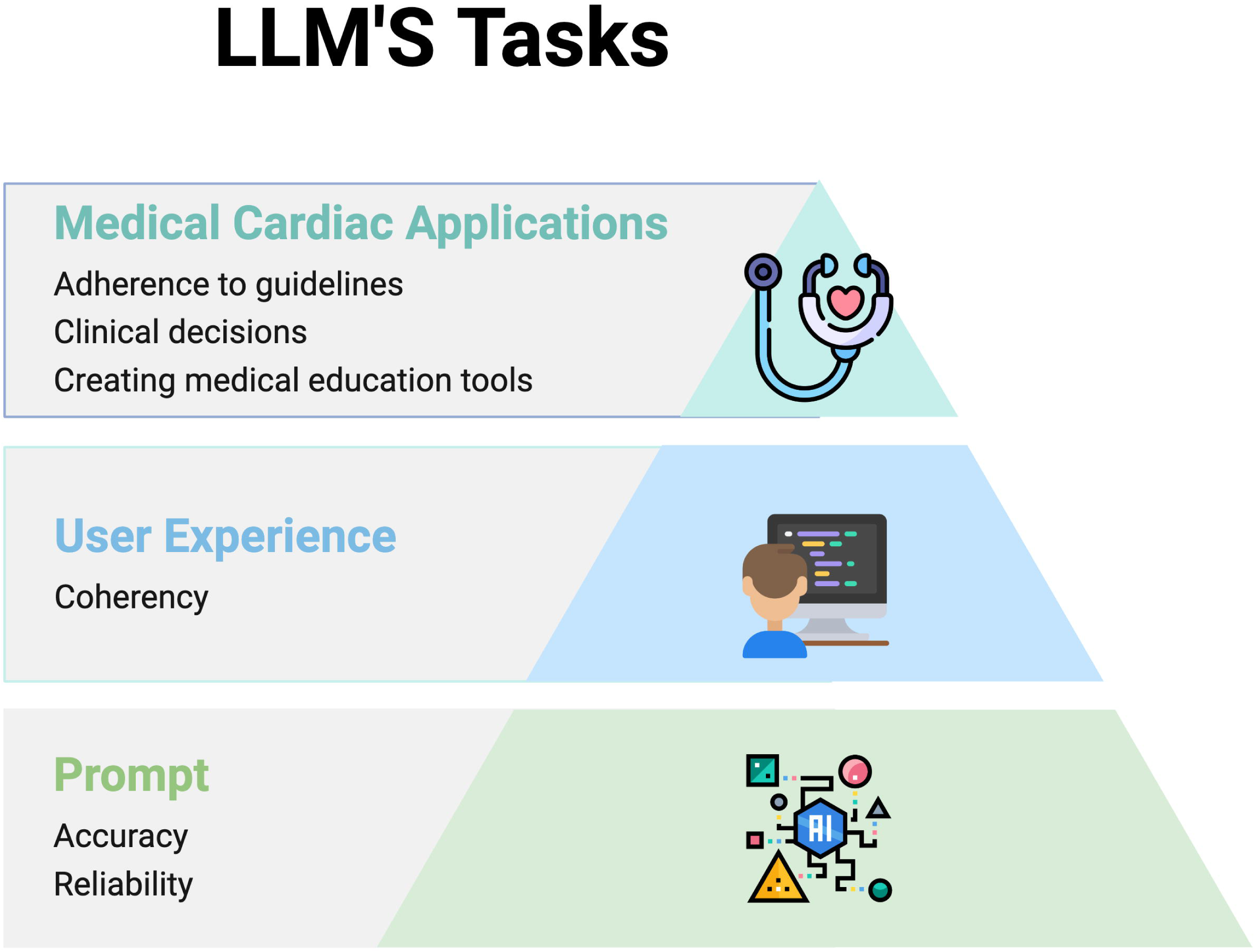
illustrates the three key aspects that the articles focus on regarding LLMs. The foundational level addresses the reliability and accuracy of LLMs, particularly in comparison to human specialists. The intermediate level concerns the interaction between the LLM and the user, emphasizing user experience. The top level highlights the specific applications of LLMs within the cardiology field.

## Discussion

The profiled studies revealed that LLMs have shown potential in various areas of cardiac management. For chronic heart conditions, LLMs like ChatGPT can accurately answer common patient questions and make educational materials easier to read^19^. This can help patient education and engagement, which is important for the management of long-term conditions. In cardiac emergencies, LLMs provided useful medical advice that people find clear, relevant, and accurate^25,26^. This indicates that LLMs can support both professionals and laypeople during critical situations, potentially improving the immediate response to life-threatening situations. For cardiac diagnostics, multimodal LLMs have performed well in interpreting ECGs, often matching or surpassing the accuracy of human specialists^28^. This means that LLMs could assist in diagnostic processes, potentially reducing the routine workload and increasing the efficiency of clinical workflows.

Despite these benefits, there are still issues to overcome before LLMs can be widely used in cardiology. One significant challenge is the variation in the accuracy and consistency of LLM responses. While some models, like ChatGPT, have performed well, others have provided less accurate or inconsistent interpretations^23,30^. This inconsistency can make it difficult to trust these tools in clinical settings.

Integrating LLMs into real-world clinical workflows is also challenging. Making sure these models work smoothly with existing healthcare systems requires significant adjustments. For instance, integrating an LLM into an electronic health records (EHR) system could help streamline documentation by automatically generating clinical notes during patient consultations. However, making sure that LLMs can seamlessly interact with EHR systems is essential for their adoption. This integration would require thorough testing to verify that data transfer is accurate and secure, preserving the integrity of patient information while enhancing the efficiency of clinical operations.

Another issue is making sure the information provided by LLMs is readable and understandable for patients. While some progress has been made, balancing readability with accurate medical information remains a challenge^20^. Ensuring that patients of varying literacy levels can understand and benefit from LLM-generated content is important for improving patient outcomes.

Additionally, the use of LLMs raises data privacy concerns ^27^. Using LLMs involves processing large amounts of sensitive patient data, so strong safeguards are needed to protect patient privacy and comply with regulations. The reviewed articles had some prominent limitations. First, half of the studies used ChatGPT-3.5. While this model is free and accessible version, its training data only extended up to September 2021 in the time the studies were conducted. This means the model could not provide real-time information, which is critical in cardiology, a continually evolving field. Access to the most current information is important for offering patients the latest and most accurate advice. Second, the studies relied on expert evaluations rather than patient feedback, creating uncertainty about how the findings apply to real-world patient interactions. Expert evaluations, while valuable, may not fully capture patient needs. Third, the studies did not involve actual patients, raising questions about whether typical patients can communicate and engage with AI-generated responses effectively. Fourth, in the cardiac monitoring articles, data were converted into descriptive text for the LLMs to interpret. This method may not accurately reflect multimodal models’ ability to analyze visual data directly, and the written descriptions could influence the prompts. Today’s latest versions of LLMs, such as ChatGPT-4o, can analyze information directly from images, offering a more accurate assessment of their capabilities to interpret visual data.

This review has several limitations. First, it included only a small number of articles, which limits the breadth of data analyzed. Second, the diversity of tasks and methods within the studies prevented a meta-analysis. Third, due to the rapid advancements in generative AI, newer applications and studies might have emerged since completing this review. Fourth, most included studies were conducted in high-resource settings, potentially limiting the generalizability of the findings to environments with fewer resources. Finally, most studies present in-silico results; for real-world adoption, prospective clinical trials are necessary.

In conclusion, LLMs demonstrate considerable potential in the field of cardiology, particularly in educational applications and routine diagnostics. However, their performance remains inconsistent across various clinical scenarios, particularly in acute care settings where precision is critical. Enhancing their accuracy in interpreting real-world complex medical data and emergency response guidance is essential before integration into clinical practice.

## Supporting information

Material 1. Literature search strategy, Table 1. Quality Assessment of Diagnostic Accuracy Studies-2

## Data Availability

All data produced in the present work are contained in the manuscript.

https://pubmed.ncbi.nlm.nih.gov/38451243/

https://pubmed.ncbi.nlm.nih.gov/37832625/

https://pubmed.ncbi.nlm.nih.gov/38194058/

https://pubmed.ncbi.nlm.nih.gov/38127304/

https://pubmed.ncbi.nlm.nih.gov/37094282/

https://pubmed.ncbi.nlm.nih.gov/37640177/

https://pubmed.ncbi.nlm.nih.gov/38081504/

https://pubmed.ncbi.nlm.nih.gov/38481520/

https://pubmed.ncbi.nlm.nih.gov/38348835/

