## Supplementary material for "Large Language Models in Cardiology: A Systematic Review": Material 1. Literature search strategy, Table 1. Quality Assessment of Diagnostic Accuracy Studies-2

### Supplementary Online Content

**Supplementary Material 1.** Literature search strategy…………..……………..…..…..… 2

**Supplementary Table 1**. Quality Assessment of Diagnostic Accuracy Studies-2......…..3

**References**…………………………………………………………………………..……….…..4

This supplementary material has been provided by the authors to give readers additional information about the work.

#### Supplementary Material 1: Literature search strategy

Database: Ovid MEDLINE(R) and Epub Ahead of Print, In-Process & Other Non-Indexed Citations and Daily <1946 to February 14, 2021>
Search Strategy:
--------------------------------------------------------------------------------
1 ((Echocardiography) OR (Arrhythmias) OR (Cardiac Output) OR (Heart Failure) OR (Heart Valve Diseases) OR (Myocardial Ischemia) OR (acute coronary syndrome)) OR (electrocardiogram) OR (EKG) OR (ECG) OR (Aortic stenosis)).
2 ((ChatGPT) OR (large language models) OR (OpenAI) OR (Microsoft bing) OR (google bard) OR (google gemini)).
3 1 and 2

***************************

**Supplementary Table 1: Quality Assessment of Diagnostic Accuracy Studies-2 (QUADS-2) risk of bias assessment**.

Abbreviations: Pt. patient; Ref. reference. ✗ = high risk of bias; ✓ = low risk of bias.

[a]Failing to describe their study population; [b]external validation; [c]Who performed the annotations? [d]Did all patients receive the same reference standard? Were all patients included in the analysis? [e]Failing to specify ethical approval.

| | Risk of bias | | | | |
|---|---|---|---|---|---|
| **Author** | **Pt. selection[a]** | **Index test[b]** | **Ref. standard[c]** | **Flow and timing[d]** | **Data management[e]** |
| Dimitriadis, Fotis et al.[1] | ✓ | ✓ | ✓ | ✓ | ✓ |
| Riddell, Craig W et al.[2] | ✗ | ✗ | ✓ | ✗ | ✓ |
| Krittanawong, Chayakrit et al.[3] | ✓ | ✓ | ✓ | ✓ | ✓ |
| Rouhi, Armaun D et al.[4] | ✓ | ✓ | ✓ | ✓ | ✓ |
| Hillmann, Henrike A K et al.[5] | ✓ | ✓ | ✓ | ✓ | ✓ |
| Van Bulck, Liesbet, & Philip Moons[6] | ✓ | ✓ | ✗ | ✓ | ✗ |
| Birkun, A. A., & Gautam, A.[7] | ✓ | ✓ | ✓ | ✓ | ✓ |
| Scquizzato, Tommaso et al.[8] | ✓ | ✓ | ✓ | ✓ | ✓ |
| Safranek, Conrad W et al.[9] | ✓ | ✓ | ✓ | ✓ | ✓ |
| Lee, Paul C et al[10] | ✓ | ✓ | ✓ | ✓ | ✓ |
| Harskamp, Ralf E, & Lukas De Clercq.[11] | ✓ | ✓ | ✓ | ✓ | ✓ |
| Günay, Serkan et al.[12] | ✓ | ✓ | ✓ | ✓ | ✓ |
| Fijačko, Nino et al.[13] | ✓ | ✓ | ✓ | ✓ | ✓ |
| Zhu, Lingxuan et al.[14] | ✓ | ✓ | ✓ | ✓ | ✓ |
| Kangiszer, Gyula et al.[15] | ✓ | ✓ | ✓ | ✓ | ✓ |
